# The impact of fiscal food policies on the macro-economy: the case of the UK Soft Drinks Industry Levy

**DOI:** 10.1101/2025.04.23.25326252

**Authors:** Henning Tarp Jensen, Marcus R. Keogh-Brown, Linda J. Cobiac, Cherry Law, Nina T. Rogers, Harry Rutter, Martin White, Richard D. Smith

**Author notes:** Corresponding author at: Department of Global Health and Development, London School of Hygiene & Tropical Medicine, 15-17 Tavistock Place, London, WC1H 9SH, United Kingdom. E-mail address (H. T. Jensen).

## Abstract

The UK government announced the ‘Soft Drinks Industry Levy’ (SDIL), a tiered tax on sugar-sweetened beverages (SSBs), in 2016 and implemented the tax in 2018. Traditional economic assessments of SSB tax interventions focus on assessing healthcare cost implications using the standard Cost-of-Illness methodology. No studies have so far attempted to apply macroeconomic assessment methods to assess net macroeconomic impacts of SSB taxes, including anticipated standard tax-efficiency losses and macroeconomic gains from reduced health externalities, or to assess trade-offs between macroeconomic and health outcomes. We established a novel framework consisting of a dynamically-recursive sectoral macroeconomic Computable General Equilibrium (CGE) model of the UK economy linked to the nutrition-focused multi-state lifetable PRIMEtime epidemiological model, and applied this framework to estimate the future macroeconomic impacts of the SDIL, and trade-offs with future health gains, over 2018-2040. Overall, the SDIL is estimated to lead to future health gains of 86,951 Disability-Adjusted Life Years (DALYs) saved, and health-related macroeconomic gains of £2.49bn, including £1.33bn due to shifting consumer demand exposures, and £1.15bn due to SSB product reformulation. Due to additional tax-efficiency losses, the SDIL is estimated to result in a limited overall cumulative real GDP cost of £2.01bn (0.004%), and in an SDIL cost-effectiveness ratio of £23,116 per DALY saved, which suggests that the SDIL is cost-effective. Since product reformulation accounts for 46% of macroeconomic gains and makes up for >25% of overall tax-efficiency losses, this demonstrates the importance of SSB tax schemes to incentivize product reformulation to ensure that they maximize health impacts and cost-effectiveness.

## 1. Introduction

Food taxes are increasingly being implemented to reduce consumption of less healthy foods (Lauer et al. 2022). Food taxes belong to the class of Pigouvian taxes and, as such, they may be used to address externalities of market transactions, and thereby attempt to align consumers’ marginal benefits with the true costs of consumption (Pigou 1920). Consumption of sugar sweetened beverages (SSBs) is associated with both negative health outcomes and a range of negative health-related fiscal externalities, due to increased prevalence of a range of non-communicable diseases (NCDs). As a result, the World Health Organization (WHO) has recommended taxes on SSBs to reduce consumption of sugar among children and adults (WHO 2022).

On 16 March 2016, the UK government announced its intention to implement a two-tier SSB tax to be levied on both manufacturers and importers. The Soft Drinks Industry Levy (SDIL) was implemented on 6 April 2018, and taxed manufacturers and importers £0.24 per litre for drinks with >8g sugar/100 mL (High Levy), £0.18 per litre for drinks with 5-8g sugar/100 mL (Low Levy), and no charge for drinks with <5g sugar/100 mL (No Levy) (HM Revenue & Customs 2016). The following types of drinks are exempt from the SDIL: 100% fruit juice, >75% milk (or a milk replacement), >1.2% alcohol (or an alcoholic beverage replacement), or drinks which have UK sales <1 million litres/year (HM Revenue & Customs 2016). The staggered announcement and implementation process, which allowed for a two-year producer reformulation period, served the twin purposes of stimulating reformulation by SSB manufacturers to reduce sugar contents in SSBs in advance of implementation, and to subsequently encourage consumers to choose alternatives without added sugars, and with the ultimate goal to lower overweight and obesity especially among UK children (Penney et al. 2023).

SSB taxes generally increase prices (Colchero et al. 2015, Caro et al. 2018, Andreyeva et al. 2022) and decrease consumption (Colchero et al. 2015, Andreyeva et al. 2022), and modelling studies suggest that SSB taxes are likely to improve health outcomes (Lin et al. 2011; Eyles et al. 2012; Redondo, Hernández-Aguado & Lumbreras 2018). Statistical and modelling studies have similarly assessed SDIL impacts and found that as prices increase (Scarborough et al. 2020), sugar consumption generally declines (Rogers et al. 2020; Pell et al. 2021), negative stock market reactions to the announcement were short-lived (Law et al., 2020a), longer-term SSB producer turnovers were mostly unchanged (Law et al. 2020b), and health outcomes generally improved (Briggs et al. 2017; Cobiac, Law & Scarborough 2022; Cobiac Law et al. 2023, Cobiac, Rogers et al.2024; Rogers, Cummins et al. 2023). However, no studies so far have studied the macroeconomic impact of SSB taxes on Gross Domestic Product (GDP) and ‘value-added’ generation, or assessed the potential net economic gains or losses (i.e. the net impact of the likely tax efficiency losses and economic gains from improved health outcomes and reduced fiscal externalities), or analysed trade-offs against health gains (i.e. the net cost per disability-adjusted life year (DALY) saved of this type of non-pharmaceutical intervention).

This study takes a whole-economy approach to estimating the full economic costs and benefits of implementing the tiered SDIL levy, including both tax efficiency losses, i.e. welfare losses related to raising UK tax revenues, and economic gains from improved health outcomes which leads to more productive workers and lower fiscal externality costs. Specifically, we assess the health and macroeconomic impacts of the SDIL using a dynamically-recursive sectoral macroeconomic model of the UK economy based on the Standard Computable General Equilibrium (CGE) model (Löfgren et al. 2002), linked to the PRIMEtime epidemiological impact model (Cobiac et. al. 2016; Cobiac et al. 2022; Cobiac, Law et al. 2023, Cobiac, Rogers et al.2024) to enable estimation of both health and overall macroeconomic impacts of the SDIL. The standard approach has traditionally been to use a micro-simulation model of household demand to simulate food risk factor changes, which have subsequently been applied as shocks to an epidemiological model and used to simulate health outcomes. Our approach improves on the standard modelling approach in several ways. First, we employ a CGE model with full demand systems (i.e. demand systems which cover all commodities in the UK economy). We therefore go beyond many micro-simulation models of household demand, which are often focused only on food demand or even subsets of food demand, and, as such, we respond to WHO’s concern that *“(O)ne important limitation in most studies is the inability to fully account for potential substitution effects that could lead to consumption of other untaxed high-calorie food items…”* (WHO 2022). In addition, our CGE model accounts for spillovers to, and feedback effects from, other sectors in the economy. Our CGE model is therefore a superior tool for simulating food (and SSB) tax-related nutritional changes to be imposed on our PRIMEtime epidemiological model. Second, by simulating health outcomes using our PRIMEtime epidemiological model, we can then estimate labour force and health outcome changes, and fiscal externalities in the form of changes to health and social care costs, which, together with the SDIL, can be applied as a combined set of shocks to our CGE model and thereby allow us to estimate the full macroeconomic impact of the SDIL. This offers an advance on the standard approach where health outcomes are evaluated and monetised at fixed prices, but where it is not possible to properly assess macroeconomic impacts in the form of GDP and generation of added value, or account for resource constraints and how health-related expansions of productive labour may endogenously change wages and prices in labour and goods markets. In sum, we apply our model framework in a staggered fashion: Firstly, we apply our macroeconomic CGE model to simulate the impact of the SDIL on nutritional exposures; Secondly, we apply the changes in nutritional exposures as shocks to the PRIMEtime epidemiological model and simulate the change in health outcomes (which in turn allows us to calculate labour force changes and health and social care cost health-related fiscal externalities); Thirdly, we apply the labour force impacts and fiscal externalities, together with the SDIL, as shocks to the macroeconomic CGE model and simulate the full macroeconomic costs and benefits of the SDIL.

## 2. Background literature

In terms of SSB tax policy design, WHO recommends a focus on five key elements: (1) taxable products, (2) tax types, (3) tax structures, (4) tax base, and (5) tax rates (WHO 2022). In terms of taxable products, the exemptions from the SDIL (e.g. fruit juices and sweetened or flavoured milk drinks) are at odds with the WHO recommendation that all categories of SSBs should be taxed (ibid.). In terms of SSB tax type, WHO recommends SSB excise taxes over SSB tiered taxes. Hence, while tiered taxes may encourage reformulation, WHO argues that this type of tax “…requires substantially higher technical tax administration capacity…” (ibid.) When considering the tiered SDIL, it is, however, noticeable that the policy exempts drinks which have UK sales <1 million litres/year. This implies that the SDIL is unlikely to introduce potentially excessive relative administrative burdens on small- and medium-sized producers.

Research has documented many pathways of SDIL impacts. These include: (1) under-shifting of the £0.24/litre high tax levy to consumer prices, equivalent to a 31% pass through rate (Scarborough et al. 2020); (2) varied sugar purchasing impacts two years after announcement (but before implementation), where impacts varied between a 5.1g/household/week increase from changed High Levy SSB purchasing (possibly from hoarding), a 4.4g/household/week reduction from changed Low Levy SSB purchases, and a 5.7g/household/week increase from changed No Levy SSB purchases (Rogers et al. 2020); (3) reductions in sugar purchases, one-year after implementation, which varied between 16.2g/household/week from changed High Levy SSB purchases, 11.5g/household/week from changed Low Levy SSB purchases, and 8.0g/household/week from changes to all SSB purchases (Rogers, Pell et al. 2023); (4) negative stock market return for four major UK-operating SSB producers on the day of SDIL announcement, but no long-term stock market impacts (Law et al. 2020a); (5) temporary post-announcement reduction in UK-operating SSB producer turnovers, but no post-implementation impacts on SSB producer turnovers (Law et al. 2020b); (6) a 1.6% decrease in prevalence of obesity among year 6 girls in English primary schools (Rogers, Cummins et al. 2023); (7) reformulation of SSB products between announcement and implementation of the SDIL scheme which led producers to reduce sales-weighted sugar contents by 0.2g/100mL for the High Levy SSB category, 1.0g/mL for the Low Levy SSB category, 0.3g/100mL for the No Levy SSB category, and 1.5g/100mL for all SSB products combined (Bandy et al. 2020); and (8) a reformulation-driven switch in the structure of soft drink brands across SSB tax brackets at the point of implementation, including a 6% reduction in the proportion of High Levy SSB category drinks, and an 8% increase in the proportion of No Levy SSB category drinks, and the structural switch continued to widen from the implementation in April 2018 until March 2020 (Luick et al. 2024).

While the estimated under-shifting for the High Levy of the SDIL scheme (Scarborough et al. 2020) is consistent with the under-shifting found in a recent meta-study of SSB tax impacts on consumer prices (Andreyeva et al. 2022), the estimated SDIL High Levy pass-through rate of 31% is significantly below the meta-study estimate of 82% pass-through for general SSB taxes (ibid.) It should, however, be noted that there was significant heterogeneity in study locations and settings in the latter study, and that a majority of underlying studies used for their meta-analyses (35 of 62) were studies of local, state-level, or regional SSB taxes (ibid.) while 11 of the national studies used for their meta-analyses were studies of the Mexican 2014 SSB tax which has been demonstrated to have a pass-through rate of close to 100% (Colchero et al. 2015), and to over-shift in some sectors possibly due to local circumstances of limited competition (WHO 2022). In fact, the SDIL High Levy pass-through rate of 31% is consistent with the high sugar-content carbonated SSB pass-through rate of 40% estimated for the 2014 Chilean tiered SSB tax (Caro et al. 2018), indicating that a more limited price pass-through is consistent with tiered tax policies with significant incentives for reformulation (WHO 2022).

In terms of broader dietary impacts of SSB taxes, it is interesting to note that the two schemes of tiered SSB taxes, including the 2018 UK SDIL and the 2018 South African Health Promotion Levy (HPL), not only followed similar patterns of announcement in 2016 and implementation in 2018, but they also seem to have had fairly similar combined reformulation and consumption impacts on taxable SSB-related sugar intakes. Hence, pre-announcement to post-implementation reductions in sugar consumption from taxable beverage purchases was estimated to be 50g/household/week (41.7% reduction) for the SDIL (Rogers, Pell et al. 2023), which compares relatively well with the reduction from 16.25g/capita/day to 10.63g/capita/day (34.6% reduction) for the HPL (Stacey et al. 2021). In terms of marginal reformulation impacts of SSB tax schemes, the only available evidence is the (seemingly unique) SDIL evidence, which suggests that the SDIL resulted in a reformulation-related sales-weighted reduction in SSB sugar contents from 4.4g/100mL in 2015 to 2.9g/100 mL in 2018 equivalent to a 34.1% reduction (Bandy et al. 2020). This evidence will be employed, below, in the modelling of the broader health and macroeconomic impacts of the SDIL.

In terms of health outcomes, there is a large body of evidence connecting sugar intakes to specific NCD outcomes including cardiovascular disease (Malik et al. 2010a; Malik, Li et al. 2019), type 2 diabetes mellitus (Malik et al. 2010a; Malik et al. 2010b), some cancers (Malik, Schulze & Hu 2006; Malik, Li et al. 2019) and obesity in children and adults (Malik, Schulze & Hu 2006; Malik et al. 2013), and there is also some (albeit weaker) evidence suggesting links from sugar intakes to coronary heart disease (CHD) (de Koning et al. 2012; Malik & Hu 2019), and other metabolic conditions (Malik et al. 2010b; Malik & Hu 2019).

There is more modest statistical evidence linking SSB tax schemes to health outcomes, and this mainly includes evidence linking SSB taxation (including the SDIL) to dental caries (Tahmassebi et al. 2006; Valenzuela et al. 2021; Rogers, Conway et al. 2023)). As a consequence, most of the evidence of SSB tax impacts on health outcomes stems from epidemiological modelling of tax-related nutritional changes in sugar intakes (Lin et al. 2011; Eyles et al. 2012; Briggs et al. 2017; Redondo, Hernández-Aguado & Lumbreras 2018). We will adopt the same approach in what follows.

Within the literature on epidemiological and decision models of SSB consumption and disease outcomes, more than half of model applications measure direct medical costs as an outcome (Alcaraz et al. 2018, Alcaraz et al. 2023, Bardach et al. 2023), but none of these Cost-of-Illness (COI) type studies attempt to value labour force impacts related to changes in clinical health outcomes. Other economic studies attempt to assess financial economy outcomes (Law et al. 2020a, 2020b) and macroeconomic outcomes using simple sector-scaling of sector-level GDP (Oxford Economics 2016), but there are, to our knowledge, no studies which have so far attempted to assess the real economy impacts of SSB interventions using a whole-economy approach.

## 3. Methods

Our combined model framework consists of a dynamic adaptation of a static macroeconomic CGE model framework (Löfgren et al. 2002) and a nutrition-focussed epidemiological model called PRIMEtime which is based on multi-state lifetable methods (Cobiac et. Al. 2016; Cobiac et al. 2022; Cobiac, Law et al. 2023, Cobiac, Rogers et al. 2024). Implementation of the 2018 SDIL is hypothesized to impact tax efficiency, but also to improve health outcomes and labour force participation, and reduce health-related fiscal externalities in the form of health and social care costs, and thereby bring associated health and economic gains. The combination of our macroeconomic and epidemiological model frameworks therefore allows us to provide an overall economic assessment which accounts for both potential negative tax efficiency losses as well as anticipated economic gains from improved health outcomes.

Our UK model framework combines macroeconomic and health components. Our dynamically recursive macroeconomic CGE model component was developed from the static IFPRI Standard Model framework (Löfgren et al. 2002) and calibrated to a 2015 UK Social Accounting Matrix (SAM), which was derived from a 2015 Supply and Use Table (SUT) for the UK (ONS 2017a) and from sector-level data derived from the GTAP10 data base (Aguiar, Chepeliev et al. 2019). The static framework was turned into a dynamically-recursive model framework by adding a set of labour and capital factor-updating equations, based on initial 2015 UK sector employment data (ONS 2019b) and initial 2015 UK capital stock and depreciation rate data (ONS 2019c). The labour factor updating equations were linked to a standard demographic model (Jensen et al. 2019), which was calibrated to the “principal projection” of the 2016 National Population Projections for the UK (ONS 2016).

Data from the Living Costs and Food Survey 2015-16 (ONS 2017b) were used to disaggregate the aggregate household into five households by income quintiles; and the same Living Costs and Food Survey data set was also used to disaggregate the demographic model population projections by household income quintile, thereby allowing us to track impacts on nutritional exposures and health outcomes, and how they link to labour factor ownership across household income quintiles.

Additional UK Manufacturers’ Sales by Product (PRODCOM) data (ONS 2019a) and data on UK household purchases of SSB commodities (Kantar 2018) were used to disaggregate the aggregate Beverages commodity into separate Sugar-Free Drink, No Levy SSB (<5g/100mL), Low Levy SSB (5-8g/100mL), and High Levy SSB (>8g/100mL) commodities. An overview of the production activity and retail commodity sector accounts in the CGE model is presented in Appendix A.

Household-specific Almost Ideal Demand Systems (AIDS) were calibrated to economy-wide UK elasticity data (Muhammad et al. 2011; Meade et al. 2014) and detailed UK food system elasticity data (Tiffin 2011) thereby allowing the household demand systems to capture substitution across all sectors (including food and beverage sectors). Once the dynamically recursive macroeconomic model was fully specified and calibrated to 2015 data, it was run forward to 2018 by targeting 2015-2018 nominal and real GDP aggregates from the World Development Indicators database (World Bank 2019); And once we had established 2018 as the base year, we used historical 1998-2018 average GDP growth rates (ibid.) for constructing our 2018-2040 counterfactual growth simulation against which we measure our SDIL simulations. All simulations employed neoclassical price-clearing of goods and factor markets, and exchange rate-clearing of the current account of the balance of payments, and they all employed a savings-driven investment closure. In terms of government spending and finances, the counterfactual simulation employed a balanced macro-closure where government consumption was fixed as a proportion of final demand, while the policy simulations employed a revenue-neutral government budget closure, where government consumption was fixed at the counterfactual growth path and adjusted for changes in health and social care costs derived from health modelling.

The health component of our model framework consists of the PRIMEtime model (Cobiac et. Al. 2016; Cobiac et al. 2022; Cobiac, Law et al. 2023, Cobiac, Rogers et al. 2024). PRIMEtime is an epidemiological health impact model which was developed to simulate the potential medium- to long-term health impacts of policies that affect food intake and obesity in the UK. PRIMEtime consists of a population lifetable which is linked to a series of disease models. Each disease model simulates the changing incidence, prevalence and mortality of that disease in the modelled population over time. Within each disease model, the incidence or onset of new disease can increase or decrease depending on the consumption of different foods. This effect is quantified by the population impact fraction, which is a function of the population exposure to dietary-related risk factors (e.g. daily intake of fruit, red meat, etc.) and the relative risk dose-response relationships between the risk factors and diseases. Where diseases are influenced by more than one dietary risk factor, it is assumed that the effects are multiplicative rather than additive, i.e. that the risk factors are independent of each other.

The PRIMEtime model, which is employed here, relies on a closed-population cohort. In contrast, it was considered important to rely on ONS’ open-population 2016 National Population Projections (ONS 2016) for the purposes of calibrating household-specific baseline growth paths of population demographics and labour force ownership in our CGE model over the 25 year time span of our simulations (3 years of running our model forward from the 2015 base year of our economic SUT and SAM data to the 2018 base year of our simulations, and 22 years of policy simulations 2018-40). The discrepancy between the closed-population cohort of the PRIMEtime model and the ONS population projections underlying the CGE model represents an inconsistency in our model framework. However, the inconsistency is likely to be small for the purposes of our modelling, since our 25-year time span means that new births can attain a maximum age of 25, and since the NCD illnesses we are focussing on (Table 1) have relatively long exposure lags and therefore relatively low incidence rates in younger age groups.

**Table 1.** PRIMEtime model risk factors and mapping to CGE model commodities.

| **PRIMETIME model**<br>**food risk factors** | **CGE model**<br>**Commodities<sup>1</sup>** | **Illnesses with exposure to**<br>**PRIMETIME risk factors** |
| --- | --- | --- |
| BMI |  |  |
| BP |  |  |
| Calcium |  |  |
| Fibre |  |  |
| Fruit | c02 | CHD, Stroke, Diabetes |
| Legumes | c04 | CHD |
| Milk | c15 | Colorectal cancer |
| Nuts and seeds | c05 | CHD, Diabetes |
| Processed meat | c12 | CHD, Colorectal cancer, Diabetes |
| PUFA |  |  |
| Red meat | c11 | Colorectal cancer, Diabetes |
| Seafood | c13 | CHD |
| Sodium |  |  |
| SSB | c23-c25 | CHD, Diabetes |
| TFA |  |  |
| Vegetables | c03 | CHD, Stroke |
| Whole grains | c18 | CHD, Stroke, Diabetes |

Footnotes: <sup>1</sup>Definitions of CGE model commodity codes are included in Appendix A.
Notes: BMI – Body Mass Index; PUFA – Polyunsaturated Fatty Acids; SSB – Sugar Sweetened Beverages; TFA – Total Fatty Acids; BP – Blood Pressure; CHD – Coronary Heart Disease.

The PRIMEtime model, which is employed here, allows for simulating 17 risk factors (Table 1). The available risk factors include consumption of aggregate food items (fruit, legumes, milk, nuts and seeds, processed meat, red meat, seafood, SSB, vegetables, whole grains), consumption of specific nutrients (calcium, fibre, polyunsaturated fatty acids (PUFA), sodium, trans fatty acids (TFA)), and health indicators (body mass index (BMI), blood pressure (BP)). In order to avoid double-counting, we restricted this study to only model the former 10 aggregate food consumption risk factors.

Furthermore, the aggregation of commodities in our macroeconomic CGE model (appendix A) allowed us to map the PRIMEtime food consumption risk factors to the CGE model retail commodity categories (Table 1), and thereby to establish links between simulated food demand changes from the CGE model, and food consumption risk factor inputs to the PRIMEtime epidemiological model. Finally, our application of the PRIMEtime model focussed on simulating nutritional impacts on four major illnesses, including CHD, stroke, type 2 diabetes mellitus, and colorectal cancer (food risk factor pathways are listed in Table 1). While our modelling excludes other potential illness pathways e.g. asthma, dental carries, and other CVD sub-groups apart from CHD and stroke, PrimeTime will capture population impact fraction spillovers to other illnesses (see Table 5), and more generally estimate impacts on population health (cases of disease averted, disability-adjusted life years) and health and social care costs.

In order to assess the health and economic impacts of the SDIL, we simulate our model framework in a staggered fashion (Table 2). In simulation 1, we apply the SDIL as a shock to the CGE model and simulate changes to the entire economic system. However, we focus on extracting household-specific relative %-changes to the food consumption items listed in Table 1, and for each of the five income quintile households.

**Table 2.** Staggered simulation process for SDIL.

| Simulation | Simulation Model | Shocks | Outcomes |
| --- | --- | --- | --- |
| Simulation 1 | CGE model | SDIL | **Changes in food consumption by household and commodity type**<br>Used to calculate relative changes in PRIMETIME food risk factors |
| Simulations 2-6 | PRIMETIME model | Relative food risk factor changes (%-shocks) derived from simulation 1 and calculated by household type (five income quintile households) | **Changes in age-stratified health outcomes (measured in DALYs) and health and social care costs**<br>Used to derive labour force and health and social care cost changes for CGE model shocks |
| Simulation 7 | CGE model | SDIL + labour force and health and social care cost changes derived from simulations 2-6 | **Changes in macroeconomic outcomes** |

NOTE: Simulations 2-6 and 7 were simulated in full, but also decomposed into multiple household- and reformulation-specific simulations

In the subsequent simulations 2-6 (Table 2), we use the %-changes in food consumption from simulation 1, to derive %-shocks to the PRIMEtime model food risk factors. Table 1 illustrates our mapping between the CGE model food consumption items and the PRIMEtime model food risk factors. The mapping is generally one-to-one. We were, however, unable to perfectly match the ‘Seafood’ risk factor to our CGE model consumption items, and it was therefore mapped to the food commodity which accounts for the vast majority of UK household seafood consumption, i.e. c13 (’Processed and preserved fish, crustaceans, molluscs, fruit and vegetables’) (see appendix B for further details on how we established our mapping between PRIMEtime model risk factors and CGE model retail commodities).

In addition, the SSB risk factor maps to the three SSB commodities c023-c025. The PRIMEtime model does not distinguish between the three SSB categories, and we therefore needed to produce a weighted average %-change in SSB demand, in order to establish the %-change in the aggregate SSB food risk factor for the PRIMEtime model. The aggregate nature of the SSB category, in the PRIMEtime model, represents a limitation, but the need for weighting of our three SSB categories also gives us an avenue for accounting for the impact of reformulation within our analyses. For weighting-purposes, we employed estimates of sales-weighted sugar contents for each of the three SSB categories c023-c025, both for 2015, i.e. pre-announcement 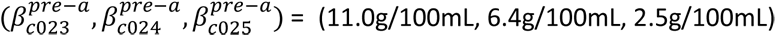 and also for 2018, i.e. pre-implementation 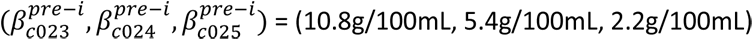 (Bandy et al. 2020). These vectors of sugar content weights can be interpreted as pre-“producer reformulation period” sugar content weights and post-“producer reformulation period” sugar content weights, and, assuming that the product reformulation, which was achieved over the producer reformulation period, remains over our 2018-2040 simulation horizon, this allows for calculating the future impact of reformulation by running separate staggered simulation assessments using each of the following separate weighting schemes:

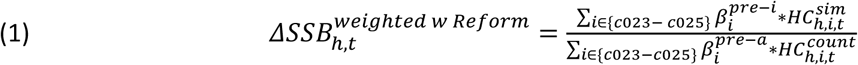

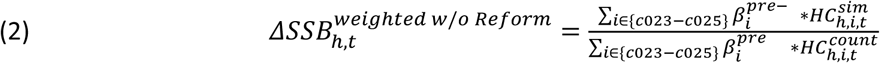

Where 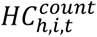 and 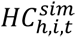 are respectively counterfactual and simulated household consumption of SSB commodity “i”, for household “h”, and in period “t”, and where 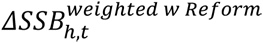 and 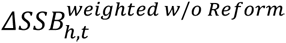 are sugar-content weighted average %-changes in the aggregate SSB food risk factor for the PRIMEtime model with and without taking account of reformulation. The impact of reformulation can then be identified as the differences of health impacts (from simulations 2-6) and economic impacts (from simulation 7) derived from duplicate simulations using the two separate *ΔSSB* measures in equations (1)-(2). This method underlies the decomposition results presented below.

We note that the PRIMEtime model is not set-up with disaggregated household types. Instead, we run separate simulations 2-6 for each of our five income quintile households, in order to simulate the health impacts of household-specific food risk factor shocks. We are thereby implicitly assuming that each of the five households have similar baseline health risk profiles, something which is probably likely to underestimate health gains and health-related fiscal externality benefits for poorer households and overestimate the gains and benefits for wealthier households.

After running simulations 2-6, we calculate our labour force shock by aggregating all age-stratified PRIMEtime DALY gains among the UK working-age population and correct for age-specific labour force participation rates. We also aggregate the PRIMEtime health and social cost changes to assess the fiscal externality change in health and social care costs. Together with the SDIL, these labour force and health-related fiscal externality shocks are subsequently imposed on the CGE model in simulation 7, where the full macroeconomic impact of the SDIL, including both tax efficiency and health-related impacts, is assessed.

## 4. Policy simulations and results

### 4.1. Health and nutritional pathways

The health-related macroeconomic gains from the SDIL amount to a combined £2.49bn, and include gains of £1.33bn due to shifting consumer demand exposures (Table 3), covering changes in household demands for all food groups and related changes in risk factors; and gains of £1.15bn due to product reformulation and reduced average sugar contents within each SSB category (Table 3), a pathway which solely affects SSB-related sugar intakes and which therefore primarily affects health outcomes from changes in CHD and Type 2 Diabetes health burdens (Table 4). In relative terms, product reformulation accounts for >46% of the total macroeconomic health-related-externality gains of the SDIL over 2018-2040. The importance of accounting for reformulation can be further illustrated by noticing that, without it, the SDIL cost-effectiveness ratio would be £36,360 per DALY saved (as opposed to our finding of an overall cost-effectiveness ratio of £23,116 per DALY saved, see next sub-section 4.2). Our approach to measuring product reformulation does not allow for assessing marginal health externality impacts for individual SSB categories. By implication, we cannot assess the overall marginal macroeconomic and health impacts of the 18p and 24p levy tiers separately.

**Table 3.**
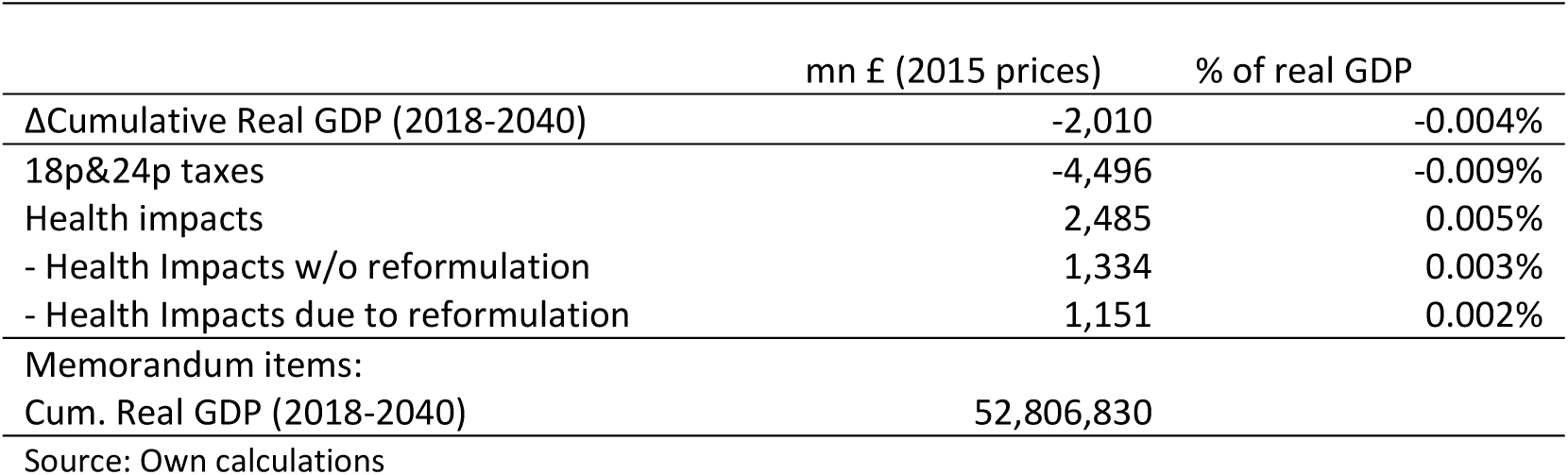
SDIL impacts (ΔCumulative real GDP; 2018-2040; mn £ ín 2015 prices)

**Table 4.** Decomposition of SSB tax and reformulation* impacts on disability-adjusted life years (DALYs) and health costs (by exposure type)

| Illness type | ΔDALYs (person-years) | | | ΔHealth Care Costs (mn £, 2015 prices) | | |
| --- | --- | --- | --- | --- | --- | --- |
|  | SDIL w/o reformulation | Reformulation* | SDIL & reformulation | SDIL w/o reformulation | Reformulation* | SDIL & reformulation |
| Fruits | -214 | 0 | -214 | 0.6 | 0.0 | 0.6 |
| legumes | 15 | 0 | 15 | -0.1 | 0.0 | -0.1 |
| milk | 236 | 0 | 236 | -0.5 | 0.0 | -0.5 |
| nuts and seeds | -400 | 0 | -400 | 1.9 | 0.0 | 1.9 |
| processed meat | 168 | 0 | 168 | -0.6 | 0.0 | -0.6 |
| red meat | 27 | 0 | 27 | -0.1 | 0.0 | -0.1 |
| seafood | 0 | 0 | 0 | 0.0 | 0.0 | 0.0 |
| sugar-sweetened beverages | 47,131 | 40,703 | 87,834 | -167.7 | -146.5 | -314.2 |
| vegetables | 18 | 0 | 18 | -0.1 | 0.0 | -0.1 |
| whole grain foods | -3 | 0 | -3 | 0.0 | 0.0 | 0.0 |
| Total | 46,978 | 40,703 | 87,681 | -166.4 | -146.5 | -312.9 |

Source: own calculations; Note: \*Reformulation measured by pre- and post-intervention avg. sugar contents from Bandy et al (2020)

When we look at the impacts on DALY health outcomes (Table 4), we see that the reformulation and direct own price effects on SSB consumption dominates. Hence, while the SDIL leads to an overall 46,978 DALY gain, reduced consumption of SSBs, by itself, leads to a gain of 47,131 DALYs indicating that spillover effects to other risk factors account for a loss of 153 DALYs. Moreover, reformulation leads to an overall 40,703 DALY gain, and this is completely dominated by gains from reduced SSB consumption. It is also noticeable that reformulation accounts for approximately 46.4% of total health outcome gains, which is on a par with reformulation accounting for 46.3% of total macroeconomic health gains.

When we add up all of the marginal health impacts of the SDIL and the added reformulation impact, we see that the total health gains amount to 86,951 DALYs gained and £312.9mn (in 2015 prices) saved on health and social care costs over 2018-2040 (Table 4). In terms of illness burdens, it is also clear from Table 5 that health gains stem mainly from reduced disease burdens of Type 2 Diabetes (70,032 DALY gain) and CHD (17,264 DALY gain), and this is also reflected in health and social care cost savings, which are dominated by savings of £203.7mn (2015 price) from a reduced Diabetes burden, and savings of £110.9mn from a reduced CHD burden.

**Table 5.** Decomposition of SSB tax and reformulation* impacts on disability-adjusted life years (DALYs) and health costs (by illness type)

| Illness type | ΔDALYs (person-years) | | | ΔHealth Care Costs (mn £, 2015 prices) | | |
| --- | --- | --- | --- | --- | --- | --- |
|  | SDIL w/o reformulation | Reformulation* | SDIL & reformulation | SDIL w/o reformulation | Reformulation* | SDIL & reformulation |
| CHD | 9,104 | 8,157 | 17,261 | -57.9 | -53.0 | -110.9 |
| stroke | -26 | 75 | 49 | 0.8 | 0.5 | 1.3 |
| diabetes, type 2 | 37,602 | 32,429 | 70,031 | -109.3 | -94.4 | -203.7 |
| breast cancer, females only | 20 | 17 | 37 | 0.2 | 0.2 | 0.4 |
| colorectal cancer | 272 | 20 | 292 | -0.3 | 0.2 | -0.1 |
| liver cancer | 1 | 1 | 1 | 0.0 | 0.0 | 0.0 |
| kidney cancer | 4 | 3 | 7 | 0.0 | 0.0 | 0.1 |
| pancreatic cancer | 1 | 1 | 2 | 0.0 | 0.0 | 0.1 |
| Total | 46,978 | 40,703 | 87,681 | -166.4 | -146.5 | -312.9 |

Source: own calculations; Notes: \*Reformulation measured by pre- and post-intervention avg. sugar contents from Bandy et al (2020)

### 4.2. Macroeconomic real GDP impact

Overall, we estimate that implementing and maintaining the SDIL will only reduce cumulative real GDP by roughly 0.004% or £2.01bn over 2018-2040 (Table 3). Over the same period of time, we estimate that the scheme will save 86,951 DALYs (Table 4), indicating that the cost-effectiveness ratio will be £23,116 per DALY saved.

A decomposition of the overall macroeconomic cost shows that overall tax-related efficiency losses, resulting from the SDIL implementation, will account for a £4.50bn loss in cumulative real GDP, while health gains will account for a combined £2.49bn gain (Table 3). The gain from reduced health externalities will therefore compensate for >55% of the overall efficiency losses from the SDIL; and product reformulation, alone, will account for £1.15bn and compensate for >25% of the overall efficiency losses.

## 5. Discussion and Conclusions

In this paper, we applied our framework, which combines a macroeconomic CGE model with the PRIMEtime epidemiological model, to assess the health and macroeconomic impacts of the tiered SDIL levy over the coming decades. Our simulations, with a focus on SDIL-related changes in nutritional exposures of 10 aggregate food consumption risk factors, resulted in SDIL health gains of 86,951 DALYs saved, and in health-related macroeconomic gains of £2.49bn, including gains of £1.33bn due to shifting consumer demand exposures, and gains of £1.15bn due to product reformulation. Not surprisingly, we also estimated that there would be an overall tax efficiency loss. Overall, we estimate a limited net cumulative real GDP loss of 0.004% or £2.01bn up to 2040.

In general, imposition of indirect taxes, such as the SDIL, creates distortions in the form of wedges between producer and consumer prices, and this, almost invariably, creates welfare losses. However, we also found that health gains were estimated to be sufficiently high so as to lower the overall cost effectiveness ratio of the SDIL scheme to be £23,116 per DALY saved. Even though the SDIL scheme is a non-pharmaceutical food tax intervention, and even though there is a fundamental methodological difference between the concepts of DALYs and QALYs, so that our cost-effectiveness ratio calculations are therefore not directly comparable to the cost-effectiveness thresholds of UK’s National Institute for Health and Care excellence (NICE), it is interesting to note that our estimated SDIL cost-effectiveness ratio per DALY saved compares well with NICE’s cost-effectiveness threshold range of £20,000-30,000 per QALY saved.

Achieving producer SSB product reformulation was the primary aim for HM Treasury in setting up the staggered announcement and implementation of the tiered SDIL. In this context, we also found evidence that UK SSB producers’ product reformulation appears to be important both in terms of achieving beneficial health impacts and reaping macroeconomic benefits from reducing negative health-related externalities. Hence reformulation is estimated to account for around 46% of both economic and health gains, and to make up for >25% of overall macroeconomic tax-efficiency losses. This illustrates that it is important, as part of SSB tax schemes, to incentivize product reformulation in order to ensure that this type of food tax scheme is cost-effective.

We have presented an approach to assessing SSB tax schemes, which is in many respects novel. The standard approach typically combines a demand-focussed microsimulation model with an epidemiological model. This allows for simulation of SSB tax induced changes to food risk factor exposures, and the epidemiological model is then able to simulate resulting health and medical cost outcomes following a standard COI-approach (Alcaraz et al. 2018, Alcaraz et al. 2023, Bardach et al. 2023). This framework has been attractive for a long time since there have been few statistical studies of the link between SSB taxes and health outcomes, something which has been blamed on long lags from exposure to illness onset (WHO 2022). Even if statistical studies of the link between SSB tax schemes and health outcomes are starting to emerge (Malik et al. 2019), there can still be value in improvements of existing simulation methods. Hence, even though our approach is also a simulation model approach, we argue that our framework improves on the standard approach in two ways. First, our framework includes the same types of demand systems as the demand system-focussed microsimulation COI-approach, but our macroeconomic framework includes full demand systems covering all food and non-food commodities in the UK economy, and it has the added advantage that it accounts for additional feedback effects across the economy which may nuance the size and scope of food consumption-focussed spillover effects. Second, our framework allows us not only to simulate SSB tax induced changes to food risk factor exposures, but it can also subsequently assess the net impact of negative tax inefficiencies and beneficial reductions in health externalities on macroeconomic indicators of interest to policymakers.

Overall, we found our novel framework to be valuable in terms of providing a balanced assessment of the health and macroeconomic impacts of the SDIL. While the estimated macroeconomic efficiency losses may seem to be at odds with earlier findings that SSB producer turnover and stock market value is unaffected by SDIL in the long-term (Law et al. 2020a, 2020b), these findings are not necessarily contradictory. Stock market value is a financial measure that is highly dependent on future expectations at a given point in time, and may not always be fully aligned with real economy business-as-usual analyses; and the impact on turnover does not necessarily need to translate one-to-one to potential changes in profitability. Furthermore, the earlier findings only referred to the four main UK SSB producers, and therefore didn’t reflect the broader SSB production sector (ibid.) In terms of price pass-through, we can also confirm that our model estimated instantaneous price pass-through of 100% in the first time period, but that price pass-through declined to 65% in the long-term. This is higher than has previously been found for the UK (Scarborough et al. 2020), but it is also lower compared to other experiences with SSB tax schemes (Colchero et al. 2015; Andreyeva et al. 2022).

In terms of limitations, we have taken a conservative approach to valuation and risk modelling. Hence, we have reported cumulative health and macroeconomic outcomes without adjusting for reduced time value of future gains and losses; we have restricted ourselves to focus on modelling of a subset of food consumption risk factors and illness pathways in the PrimeTime model; we have limited ourselves to look at health outcomes from NCDs in adults; and we have limited ourselves to model product reformulation based on reformulations achieved during the “policy reformulation period” between 2016-18, and thereby not allowed for additional long-term product reformulation beyond SDIL implementation in 2018 (Luick et al. 2024). Moreover, our epidemiological model did not allow for detailed health modelling of individual Low and High Levy SSBs, and this may have led to underestimation of health benefits for low-income households and overestimation of health benefits for high-income households; and our model framework consisted of staggered applications of two stand-alone macroeconomic and epidemiological models, which were not fully integrated, and where feedback effects may not have been fully captured. Finally, our macroeconomic model counterfactual growth path was not adjusted for the drop in UK GDP during the Covid-19 pandemic years (2020-21) and, while UK economic growth has since resumed and while the UK, in spite of annual fluctuations, seems to be back on its long-term macroeconomic growth trajectory again, our valuation of SDIL gains and losses for 2020-21 may not have been accurately measured due to reduced real wages and potentially increased consumption of SSBs during lockdowns.

In spite of these limitations, our study adds importantly to the literature by providing a novel methodological approach for assessing the broader macroeconomic consequences of SSB tax schemes including the net impact of likely tax efficiency losses and economic gains from improved health outcomes and reduced health-related fiscal externalities. As such, our study demonstrates the value of switching away from COI-based assessments and taking a macroeconomic approach to assessing future food tax policy schemes. Beyond our methodological contribution, we also provide evidence which indicates that tiered SSB tax schemes, if properly implemented, can be cost-effective health interventions. Specifically, we found that the product reformulation incentives, embodied in the SDIL, were sufficiently strong to ensure that the tiered tax scheme is indeed cost-effective. UK policymakers can therefore take comfort in the fact that the SDIL is well-worth the costs that the scheme has entailed; and policymakers from abroad, who may consider implementing future SSB tax reforms, should learn from the SDIL experience, and ensure that transition periods between tax scheme announcement and implementation, which are meant to provide product reformulation incentives, are included as an integral feature of reform designs.

## Data Availability

Most of the data underlying the model is available upon request from the authors. However some of the data underlying the model were developed from the Global Trade Analysis Project (GTAP) database which is proprietary and cannot be shared publicly. The GTAP database may be obtained from https://www.gtap.agecon.purdue.edu/databases/v10/index.aspx.

## Acknowledgements

MW and RS led on grant application; HTJ, MKB and RS led on conceptualization, study design and methods development; HTJ and MKB led on literature review, methods application, data analysis and interpreting, text drafting; LJC was involved in methods development; and all authors were involved in interpreting data and commenting on drafts, and in editing/proof reading/formatting. The work benefitted from broader discussions within the SDIL Evaluation study team, members of which are identified here: https://www.mrc-epid.cam.ac.uk/research/studies/sdil/. This research was funded by the UK National Institute for Health and Care Research (NIHR) Public Health Research Programme, Study number: 16/130/01. MW was also supported by the Medical Research Council, grant numbers MC_UU_12015/6 and MC/UU/00006/7, and the Centre for Diet and Activity Research, a UKCRC Public Health Research Centre of Excellence, for which funding from the British Heart Foundation, Cancer Research UK, the Economic and Social Research Council, the Medical Research Council, the National Institute for Health Research, and the Wellcome Trust, under the auspices of the UK Clinical Research Collaboration, is gratefully acknowledged. The funders had no role in study design, data collection and analysis, decision to publish, or preparation of the manuscript.

## Appendix A. UK CGE model sector classification

**Table A.1.** UK CGE model sector classification of production activities and retail commodities.

| Production activity | Retail commodity | Sector label |
| --- | --- | --- |
| a01 | c01 | Cereal grains |
| a02 | c02 | Fruits |
| a03 | c03 | Vegetables |
| a04 | c04 | Legumes |
| a05 | c05 | Nuts and seeds |
| a06 | c06 | Crops nec |
| a07 | c07 | Animal products |
| a08 | c08 | Products of forestry, logging and related services |
| a09 | c09 | Fish and other fishing products; aquaculture products; support services to fishing |
| a10 | c10 | Mining products |
| a11 | c11 | Meat, red |
| a12 | c12 | Other preserved meat and meat products |
| a13 | c13 | Processed and preserved fish, crustaceans, molluscs, fruit and vegetables |
| a14 | c14 | Vegetable and animal oils and fats |
| a15 | c15 | Milk |
| a16 | c16 | Other dairy products |
| a17 | c17 | Grain mill products, starches and starch products |
| a18 | c18 | Whole grain bakery and farinaceous products |
| a19 | c19 | Other bakery and farinaceous products |
| a20 | c20 | Other food products |
| a21 | c21 | Alcoholic beverages & Tobacco products |
| a22 | c22 | Soft drinks (non-SSB) |
| a23 | c23 | Soft drinks (No Levy SSB) |
| a24 | c24 | Soft drinks (Low Levy SSB) |
| a25 | c25 | Soft drinks (High Levy SSB) |
| a26 | c26 | Textile and leather products |
| a27 | c27 | Other manufacturing |
| a28 | c28 | Utilities |
| a29 | c29 | Construction |
| a30 | c30 | Trade |
| a31 | c31 | Transport |
| a32 | c32 | Public administration |
| a33 | c33 | Education services |
| a34 | c34 | Health services |
| a35 | c35 | Recreation services |
| a36 | c36 | Other services |

Note: Classification based on own aggregation and disaggregation of original 2015 UK Supply-Use Table data (ONS 2017a).

## Appendix B. Mapping of PRIMEtime nutritional risk factors to CGE model food commodities

**Table B.1.** Mapping of PRIMEtime model food risk factors and UK CGE model retail commodities.

| PRIMETIME model food risk factor | CGE model retail commodity | CGE model retail commodity label |
| --- | --- | --- |
|  | c01 | Cereal grains |
| Fruit | c02 | Fruits |
| Vegetables | c03 | Vegetables |
| Legumes | c04 | Legumes |
| Nuts and seeds | c05 | Nuts and seeds |
|  | c06 | Crops nec |
|  | c07 | Animal products |
|  | c08 | Products of forestry, logging and related services |
|  | c09 | Fish and other fishing products; aquaculture products; support services to fishing |
|  | c10 | Mining products |
| Red meat | c11 | Meat, red |
| Processed meat | c12 | Other preserved meat and meat products |
| Seafood | c13 | Processed and preserved fish, crustaceans, molluscs, fruit and vegetables |
|  | c14 | Vegetable and animal oils and fats |
| Milk | c15 | Milk |
|  | c16 | Other dairy products |
|  | c17 | Grain mill products, starches and starch products |
| Whole grains | c18 | Whole grain bakery and farinaceous products |
|  | c19 | Other bakery and farinaceous products |
|  | c20 | Other food products |
|  | c21 | Alcoholic beverages and Tobacco products |
|  | c22 | Soft drinks (non-SSB) |
| SSB | c23 | Soft drinks (No Levy SSB) |
| SSB | c24 | Soft drinks (Low Levy SSB) |
| SSB | c25 | Soft drinks (High Levy SSB) |
|  | c26 | Textile and leather products |
|  | c27 | Other manufacturing |
|  | c28 | Utilities |
|  | c29 | Construction |
|  | c30 | Trade |
|  | c31 | Transport |
|  | c32 | Public administration |
|  | c33 | Education services |
|  | c34 | Health services |
|  | c35 | Recreation services |
|  | c36 | Other services |

Note: CGE model retail commodity classification based on own aggregation and disaggregation of original 2015 UK Supply-Use Table data (ONS 2017a).

Our mapping between food risk factors from the PRIMEtime model, and retail commodities from the CGE model is presented in Table B.1. The motivation for the chosen mapping is as follows:

- <u>Risk factor ‘Fruit’ is mapped to retail commodity c02 (‘Fruits’):</u> Household demand for fresh fruits is captured by commodity c02 (‘Fruits’) while demand for processed fruits is captured by the aggregate commodity c13 (‘Processed and preserved fish, crustaceans, molluscs, fruit and vegetables’). Since the risk factor ‘Fruit’ refers to consumption of fresh fruit, this mapping is one-to-one.
- <u>Risk factor ‘Vegetables’ mapping to retail commodity c03 (‘Vegetables’):</u> Household demand for fresh vegetables is captured by commodity c03 (‘Vegetables’) while demand for processed fruits is captured by the aggregate commodity c13 (‘Processed and preserved fish, crustaceans, molluscs, fruit and vegetables’). Since the risk factor ‘Vegetables’ refers to consumption of fresh vegetables, this mapping is one-to-one.
- <u>Risk factor ‘Legumes’ mapping to retail commodity c04 (‘Legumes’):</u> Household demand for legumes is only captured by commodity c04 (‘Legumes’), and this mapping is therefore one-to-one.
- <u>Risk factor ‘Nuts and seeds’ mapping to retail commodity c05 (‘Nuts and seeds’):</u> Household demand for nuts and seeds is only captured by commodity c05 (‘Nuts and seeds’), and this mapping is therefore one-to-one.
- <u>Risk factor ‘Red meat’ mapping to retail commodity c11 (‘Meat, red’):</u> Household demand for red meat is only captured by commodity c11 (‘Red meat’), and this mapping is therefore one-to-one.
- <u>Risk factor ‘Processed meat’ mapping to retail commodity c12 (‘Other preserved meat and</u> <u>meat products’):</u> Household demand for processed meat is only captured by commodity c12 (‘Other preserved meat and meat products’), and this mapping is therefore one-to-one.
- <u>Risk factor ‘Red meat’ mapping to retail commodity c11 (‘Meat, red’):</u> Household demand for red meat is only captured by commodity c11 (‘Red meat’), and this mapping is therefore one-to-one.
- <u>Risk factor ‘Processed meat’ mapping to retail commodity c12 (‘Other preserved meat and</u> <u>meat products’):</u> Household demand for processed meat is only captured by commodity c12 (‘Other preserved meat and meat products’’), and this mapping is therefore one-to-one.
- <u>Risk factor ‘Seafood’ mapping to retail commodity c13 (‘Processed and preserved fish,</u> <u>crustaceans, molluscs, fruit and vegetables):</u> Household demand for Seafood is overwhelmingly captured by commodity c13 (‘Processed and preserved fish, crustaceans, molluscs, fruit and vegetables’). A smaller part may be captured by the primary commodity c09 (‘Fish and other fishing products; aquaculture products; support services to fishing’). However, since household demand for c13 is roughly 21 times the size of household demand for c09, and since it was not possible to determine disaggregate household consumption of c09 into seafood and other aquaculture-related products, we chose to map the risk factor ‘Seafood’ solely to commodity c13. We also note that the c13 is a composite of processed seafood and processed fruit and vegetables, and we therefore, in our analyses, assume that an x% change in household demand for commodity c13 results in an x% change in demand for both processed seafood and for processed fruit and vegetables.
- <u>Risk factor ‘Milk’ is mapped to retail commodity c15 (‘Milk’):</u> Household demand for Milk is only captured by commodity c15 (‘Milk’), and this mapping is therefore one-to-one.
- <u>Risk factor ‘Whole grains’ is mapped to retail commodity c18 (‘Whole grain bakery and</u> <u>farinaceous products’):</u> Household demand for Whole grains is only captured by commodity c15 (‘<u>Whole grain bakery and farinaceous products</u>’), and this mapping is therefore one-to-one.
- <u>Risk factor ‘SSB’ is mapped to retail commodities c23-c25 (‘Soft drinks (No Levy SSB)’, ‘Soft</u> <u>drinks (Low Levy SSB)’, ‘Soft drinks (High Levy SSB)’):</u> Since household demand for SSBs is captured by three commodities c23 (‘Soft drinks (No Levy SSB)’), c24 (‘Soft drinks (Low Levy SSB)’), and c25 (‘Soft drinks (High Levy SSB)’), this mapping is not one-to-one. As a consequence, we weighted the %-changes in household demand for the three retail commodities in order to determine how SDIL impacted the aggregate SSB risk factor. The weighting schemes, which we employed, are described in the main body text.

